# TROMBIX-DZ: A real-world, prospective, observational study of Algerian patients with atrial fibrillation treated with rivaroxaban

**DOI:** 10.64898/2026.05.26.26353979

**Authors:** Ahlem Sarah Moulay Brahim, Samia Lekkam, Sarah Helal, Meriem Aouchar, Ismahane Benbitour, Lynda Noual, Yazid Aoudia, Naima Adjeroud, Mohamed Seddik Ait Messaoudene, Mohamed Afif, Hadj Mohammed Ali Lahmer, Houssam Eid, Nadia Laredj, Bassem Aouiche, Ryad Hamdi, Mohamed Fayçal Beddai, Samir Berboucha, Toufik Boudjelal, Samir Boumaaza, Tarik Fernane, Aldjia Kachenoura, Zaki Kaiter, Nabil Nemmar, Nadjib Lassakeur, Mohamed Mouffok, Nassima Nassour, Ghoussoun Sebbagh, Ridha Okbi

## Abstract

**Background:** Atrial Fibrillation (AF) is the most prevalent cardiac arrhythmia worldwide, representing the primary cardiac etiology of stroke. In recent years, direct oral anticoagulants (DOACs) have shown favorable results in terms of efficacy and safety in the prevention of thromboembolism in patients with AF.

TROMBIX-DZ study investigated the safety and efficacy of rivaroxaban in routine clinical settings in response to the need for real-world evidence on the use of DOACs.

**Methods:** We carried a national, multicenter, prospective, observational cohort study to evaluate the safety and efficacy of rivaroxaban in Algerian’s patients with atrial fibrillation. Patients were followed-up at 3 months intervals for 1 year. The primary outcome of this study was to evaluate the safety of rivaroxaban, reported as the frequency of treatment-emergent serious adverse events (SAEs); Secondary outcomes assessed the frequency of thromboembolic events, adverse events (AEs), and treatment persistence.

**Results:** TROMBIX-DZ enrolled 398 eligible patients with AF from 19 specialized public and private cardiology centers across different regions in Algeria. The mean age was 70.5 ± 11.94. 71.9% of patients received once daily rivaroxaban 20mg, and 28.1% received the 15mg dose. The most common comorbidities included, hypertension (77.1%), diabetes (28.6%) and heart failure (25.4%), prior strokes, TIA (8.8%) and prior major bleeding (3.1%). The mean CHA_2_DS_2_-VASc score was 3.147± 1.3, and the mean HAS-BLED score was 1.682±1.198; 14.06% of patients had Creatinine clearance < 50 ml/min.

A total of 5.77% had treatment-emergent AE, and 1.76% had treatment-emergent hemorrhagic SAE. The incidence rate (events per 100 patient-years) of treatment-emergent major bleeding events, treatment-emergent thromboembolic events and all-cause death during the study period were 2.1, 0.9, and 4.18, respectively. Treatment persistence was 75,88% at the end of the study.

**Conclusion:** TROMBIX-DZ study, the first cohort in the Maghreb region, provides important insights into the safety and efficacy of rivaroxaban in Algerian population with atrial fibrillation receiving standard medical care. Rates of major bleeding and stroke were low and broadly consistent with previous international real-world registries.

**Trial registration number:** Clinicaltrial.gov: (NCT06184204).

## Introduction

Atrial Fibrillation (AF) is the most prevalent cardiac arrhythmias, with a rising trend globally due to several factors such as an aging population **(1)**. AF is affecting around 52.6 million patients worldwide and increasing significantly in the Middle East and North Africa (MENA) region **(2)**. As of 2019, the European Society of Cardiology estimates that AF affects approximately 0.49% of the Algerian population **(3)**.

International guidelines recommend the use of Direct Oral Anticoagulants (DOACs) over Vitamin K antagonists (VKAs) to prevent ischemic stroke and thromboembolism in patients with AF, as supported by several phase III studies, demonstrating better efficacy and improving safety through reduced risk of major bleeding **(1, 4)**. Nevertheless, VKAs continue to be prescribed due to economic reasons **(5)**. In Algeria, over the recent years, a progressive shift in clinical practice toward DOACs has been observed, particularly with the introduction of the first generic of rivaroxaban “Trombix®” developed by the Algerian pharmaceutical company “Laboratoires BEKER®” into the Algerian market.

Observational studies conducted in several nations have demonstrated the safety and efficacy of rivaroxaban in real-world settings **(6, 7, 8, 9, 10)**. This type of studies is essential to complement data from clinical trials by evaluating the efficacy and safety of DOACs in more heterogeneous populations with multiple comorbidities and exposed to different treatments. Therefore, due to the scarcity of local data about DOACs utilization in AF patients in Algeria, we conducted the first prospective observational study TROMBIX-DZ involving Algerian patients with AF receiving rivaroxaban. This observational study aimed to assess the safety and efficacy of Rivaroxaban, to provide further data in Algerian population.

## Methods

We conducted a national, multicenter, prospective, observational cohort study, to evaluate the safety and efficacy of Trombix® (Rivaroxaban) in patients with atrial fibrillation.

A total of 398 consenting patients, aged ≥ 19 years, with atrial fibrillation requiring treatment with rivaroxaban and patients needed substitution of VKA therapy (labile INR or those seeking a more manageable treatment option) were enrolled from 19 specialized public and private cardiology centers across different regions in the country; East (176 patients), Center (108 patients) and West (114 patients) between December 2023 to march 2025. Patients who met one of the following criteria were excluded: contraindications to Rivaroxaban, end-stage renal disease, participation in another clinical study within 12 weeks prior to inclusion, and patients whom the investigator judge’s incapable of participating in the study.

According to the treating physician’s discretion, patients were administered a dose of 15 mg or 20 mg rivaroxaban once daily orally; a dose of 15 mg was recommended for patients with CrCl< 50 mL/min.

Patients were followed up for a period of one year. After the inclusion visit, follow-up data collection was every 3 months thereafter. The observational period for patients who discontinued therapy before the end of the study ended 30 days after the last dose of rivaroxaban.

The primary outcome of this study was to evaluate the safety of rivaroxaban, reported as the frequency of serious adverse events (SAEs), (was considered as a SAE, any event that may result in hospitalization, death, congenital anomaly, permanent disability or any event that may jeopardize the patient’s life or functional prognosis), this included major bleeding events (defined as any bleeding meeting hemodynamic criteria, requiring hospitalization with transfusion depending on the site of hemorrhage i.e. gastrointestinal bleeding, critical organ bleeding, intracranial hemorrhage, and fatal bleeding).

Secondary outcomes assessed the frequency of thromboembolic events (ischemic stroke/ transient ischemic attack (TIA)/ Myocardial Infarction (MI) / non-Central Nervous System non-CNS Systemic embolism), adverse events counting non-major bleeding events related to rivaroxaban occurring during the study period, and treatment persistence.

Baseline socio‐demographic, clinical and laboratory data including previous medical history and comorbidities, anticoagulant treatments and other concomitant medications, kidney function (CrCl) were collected at the first visit, recorded in a case report form (CRF) and verified against medical records at each visit by the study investigator. Physicians reported any serious adverse events, including major bleeding, stroke, TIA, or any other adverse event, noting yes/no responses for each event. Non-major bleeding events were reported by the patients using a specific designed cards and collected during the periodic visits. CHA_2_DS_2_-VASc and HAS‐BLED scores were calculated for each study patient to assess thrombotic and bleeding risk.

The study was conducted following approval of the Ethical Committee (EC) of Issad Hassani University Hospital of Beni Messous (N° 8 / CE/2023) and the Algerian Ministry of Industry and Pharmaceutical Production (MIPP) (012/BEK/OBS/MED/2023) in November 2023, according to the Declaration of Helsinki, Good Clinical Practice (ICH GCP E6) and current Algerian regulations. The study protocol of TROMBIX-DZ is registered in clinicaltrial.gov (NCT06184204).

the sponsor was responsible for protocol development and oversaw the design, the monitoring, the data collection and analysis. The study protocol was validated by the scientific committee of the study. The operational conduct of this study was outsourced to an independent Contract Research Organization (CRO), “Clinica Group CG.” This organization was responsible for protocol and clinical report writing, site monitoring, data collection, and management to ensure adherence to the protocol and (GCP) guidelines. The sponsor and the CRO had access to all patient’s data. Statistical analyses were carried out by an independent statistician.

### Statistical analysis

The study aimed to recruit patients with atrial fibrillation. The required sample size was calculated with estimating the incidence rate of serious AEs to be 17.7% **(8)**. Based on this percentage, a type I error risk of α = 5%, and a precision level of 4%, the calculations yielded a minimum sample size of n = 350 patients. Taking into account patients lost to follow-up, estimated around 14%, the determined sample size was n = 400 patients.

Statistical analyses of the events were descriptive and exploratory, relying on frequency tables, curves, and summary statistics (e.g., mean ± standard deviation [SD]).

Both raw incidence proportions (patients with events/number of treated patients) and incidence rates (events per 100 patient-years) are reported, with corresponding 95% confidence intervals. All statistical analyses were performed using R version 4.4.1.

## Results

### Study population

A total of 398 eligible patients with AF (72% recently diagnosed) were enrolled in this study of whom 302 (75.88%) completed all the visits. The mean duration of the study was 307.22 days (SD= 120.93; median=360). The majority of patients (71.9%) received rivaroxaban 20mg, and the remaining (28.1%) received the 15mg dose (**Fig. 1)**. A total of 253 were VKA naïve, 105 were on VKA at inclusion and 36 had a history of VKA use.

**Figure 1.**
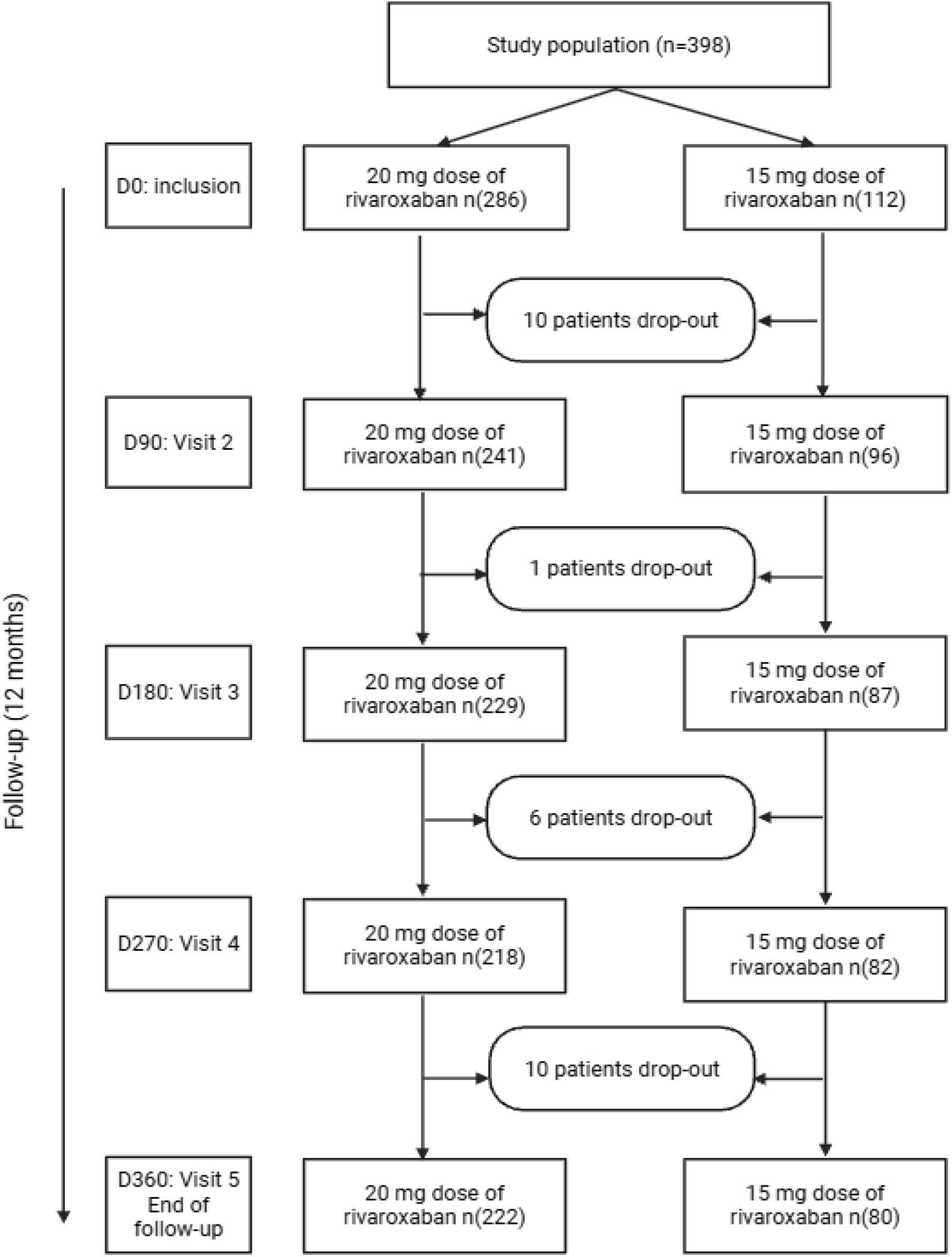
Patient disposition during the study.

The baseline demographic and clinical features of patients are reported in **Table 1**. Patients’ mean age was 70.5 ± 11.94; 227 (57%) of all patients were aged ≥70, and 209 (52.5%) were female with a sex ratio of M/F of 0.9. The most common comorbidities were, hypertension (77.1%), diabetes (28.6%) and heart failure (25.4%). Overall, 8.8% of patients had experienced thromboembolic events (strokes and TIA), and 3.1% had experienced major bleeding. The mean CHA_2_DS_2_-VASc score was 3.147 ± 1.3, and the mean HAS-BLED score was 1.682 ± 1.198. CrCl values were reported for the majority of the patients (73.6%); 14.06% had CrCl < 50 ml/min.

**Table 1.**
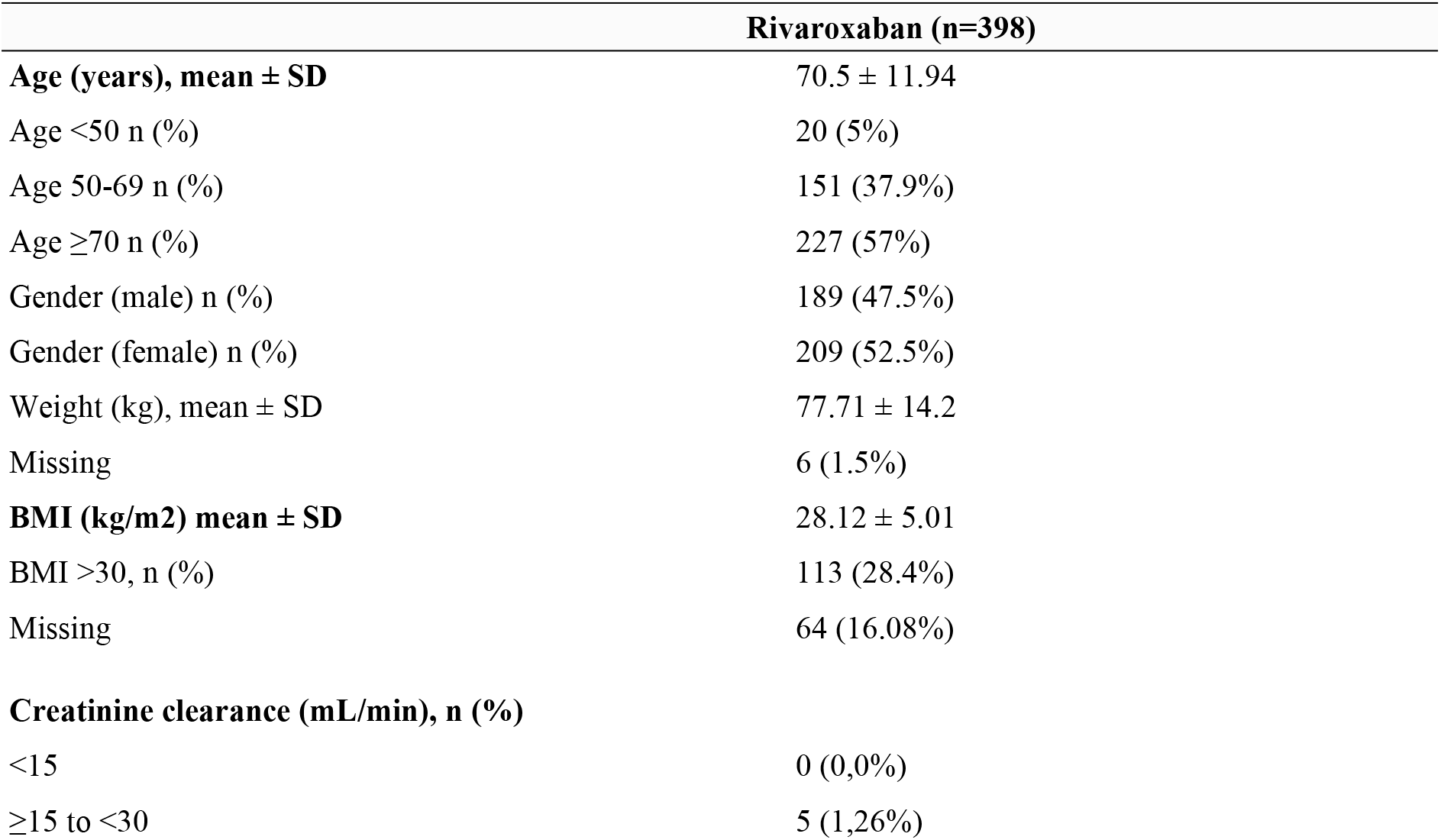
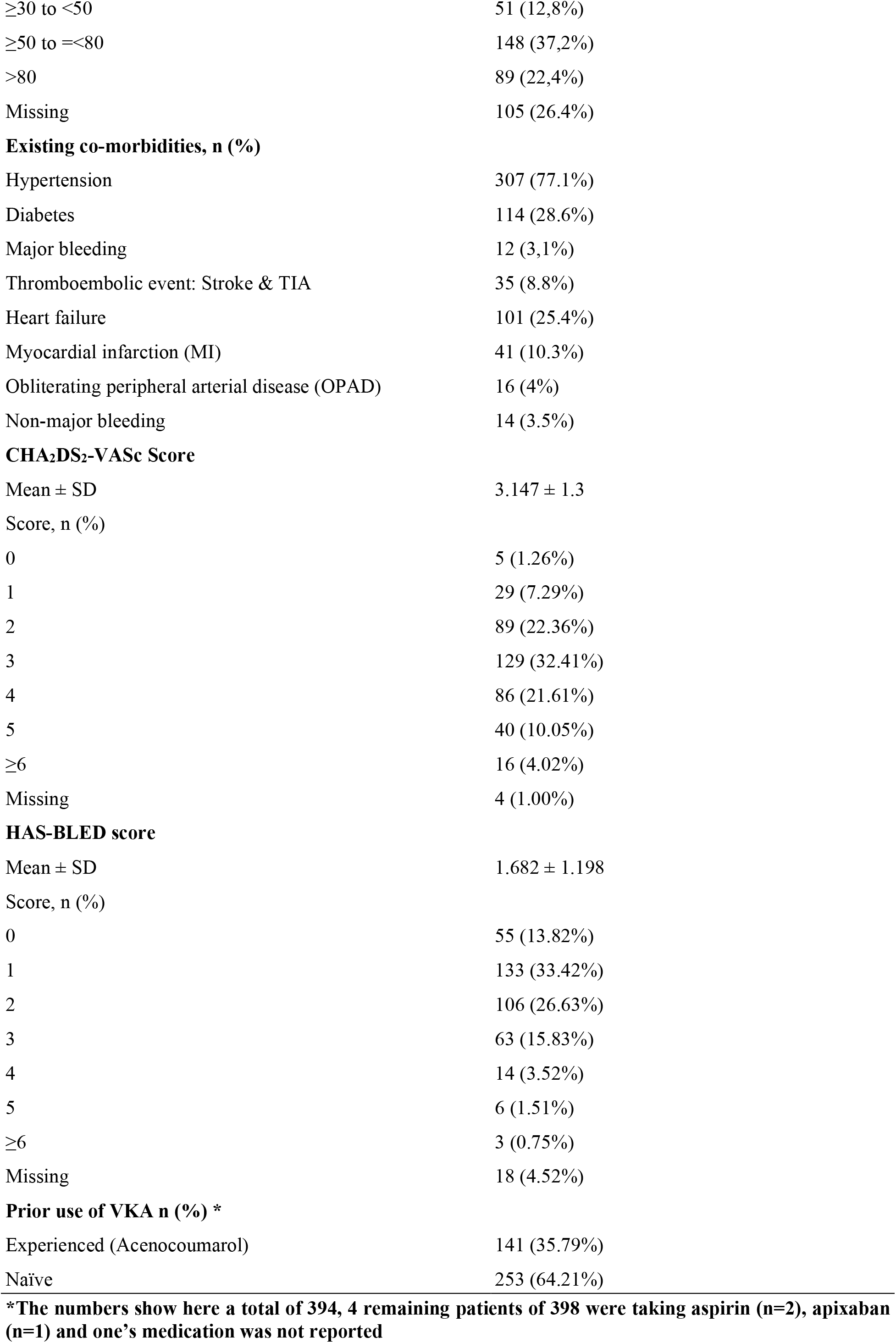
Baseline demographics and clinical characteristics of patients.

### Outcomes

A total of 23 patients (5.77%) had treatment-emergent AE, and 7 (1.76%) had treatment-emergent hemorrhagic SAE.

There were 7 treatment-emergent major bleeding events in 7 patients, aged >65, with HAS-BLED score≥2 and CHA_2_DS_2_-VASc≥4, the incident rate was 2.1 events per 100 patient-years **(Table 2)**. In three patients, HAS-BLED score was equal to 3 indicating a high risk of major bleeding.

**Table 2.** Treatment-emergent thromboembolic and bleeding events and all cause death.

|  | **Rivaroxaban (N=398)** |  |
| --- | --- | --- |
|  | **Incidence proportion n (%)** | **Incidence rate. events per 100 patient-year (95%CI)** |
| Thromboembolic event (Stroke and TIA) | 3 (0.75%) | 0.9 events per 100 patient-years [0.18 ; 2.62] |
| TIA | 1 (0.25%) | 0.3 events per 100 patient-years [0.01 ; 1.66] |
| Stroke | 2 (0.5%) | 0.6 events per 100 patient-years [0.07 ; 2.16] |
| Major bleeding | 7 (1.76%) | 2.1 events per 100 patient-years [0.85 ; 4.33] |
| Intracranial hemorrhages | 1 (0.25%) | 0.3 events per 100 patient-years [0.01 ; 1.66] |
| Gastrointestinal hemorrhages | 4 (1.01%) | 1.2 events per 100 patient-years [0.33 ; 3.07] |
| Hemorrhages requiring hospitalization with transfusion | 2 (0.5%) | 0.6 events per 100 patient-years [0.07 ; 2.16] |
| Bleeding of vital organs | 1 (0.25%) | 0.3 events per 100 patient-years [0.01 ; 1.66] |
| Fatal hemorrhages | 1 (0.25%) | 0.3 events per 100 patient-years [0.01 ; 1.66] |
| Non-major bleeding events | 14 (3.52%) | 4.25 events per 100 patient-years [2.32 - 7.13] |

The incidence rate of major gastrointestinal bleeding was 1.2 events per 100 patient-years; bleeding requiring hospitalization with transfusion occurred at a rate of 0.6 events per 100 patient-years, and the incidence rate of critical organ bleeding was 0.3 events per 100 patient-years, including intracranial hemorrhage. The incidence rate of fatal bleeding (intracranial hemorrhage) was 0.3 events per 100 patient-years with HAS-BLED score of 3.

There were 3 treatment-emergent thromboembolic events in 3 patients (0.9 events per 100 patient-years), stroke for 2 patients (0.6 events per patient-year), and a transient ischemic attack for the third (0.3 events per 100 patient-years) **(Table 2)**.

Treatment emergent AEs occurred in 5.77% of patients with incidence rate of 7.02 events per 100 patient-years. Fourteen patients experienced non-major bleeding with an incidence rate of 4.25 per 100 patient-years.

The overall numbers of thromboembolic events (n=3), major bleeding (n=7) and all-cause death (n=14) were low and increased slightly over time **(Fig. 2A)**. All-cause death rate was 4.18 events per 100 patient-years during the study period, with one treatment-emergent cause of death attributed to intracranial hemorrhage. other causes included: sudden death (n=3), heart failure (n=1), uterine cancer (n=1) and Unknown cause of death (n=8).

**Figure 2A.**
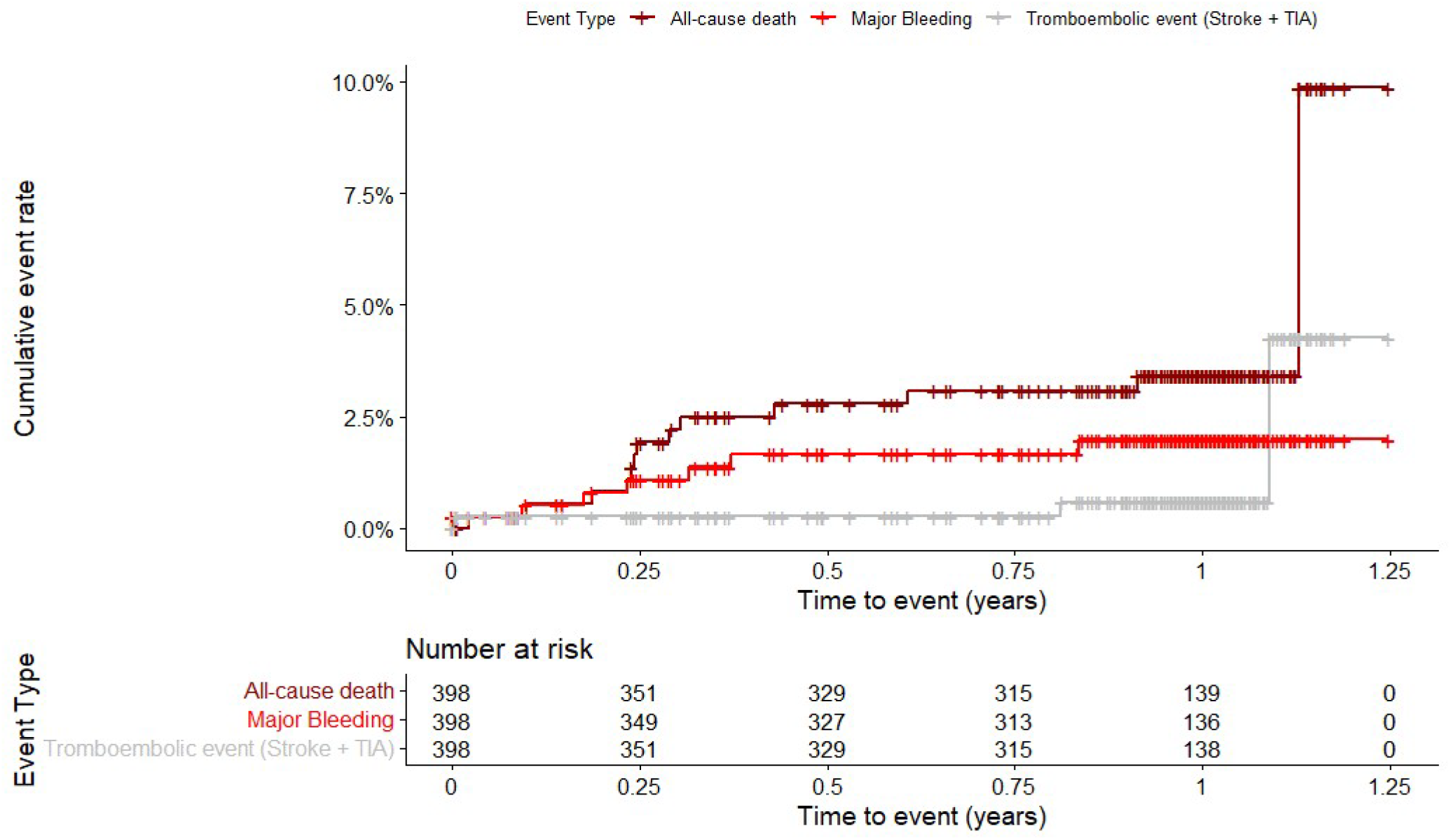
Cumulative rates (Kaplan-Meier) for all-cause death, major bleeding events, and thromboembolic events (stroke/TIA).

**Figure 2B.**
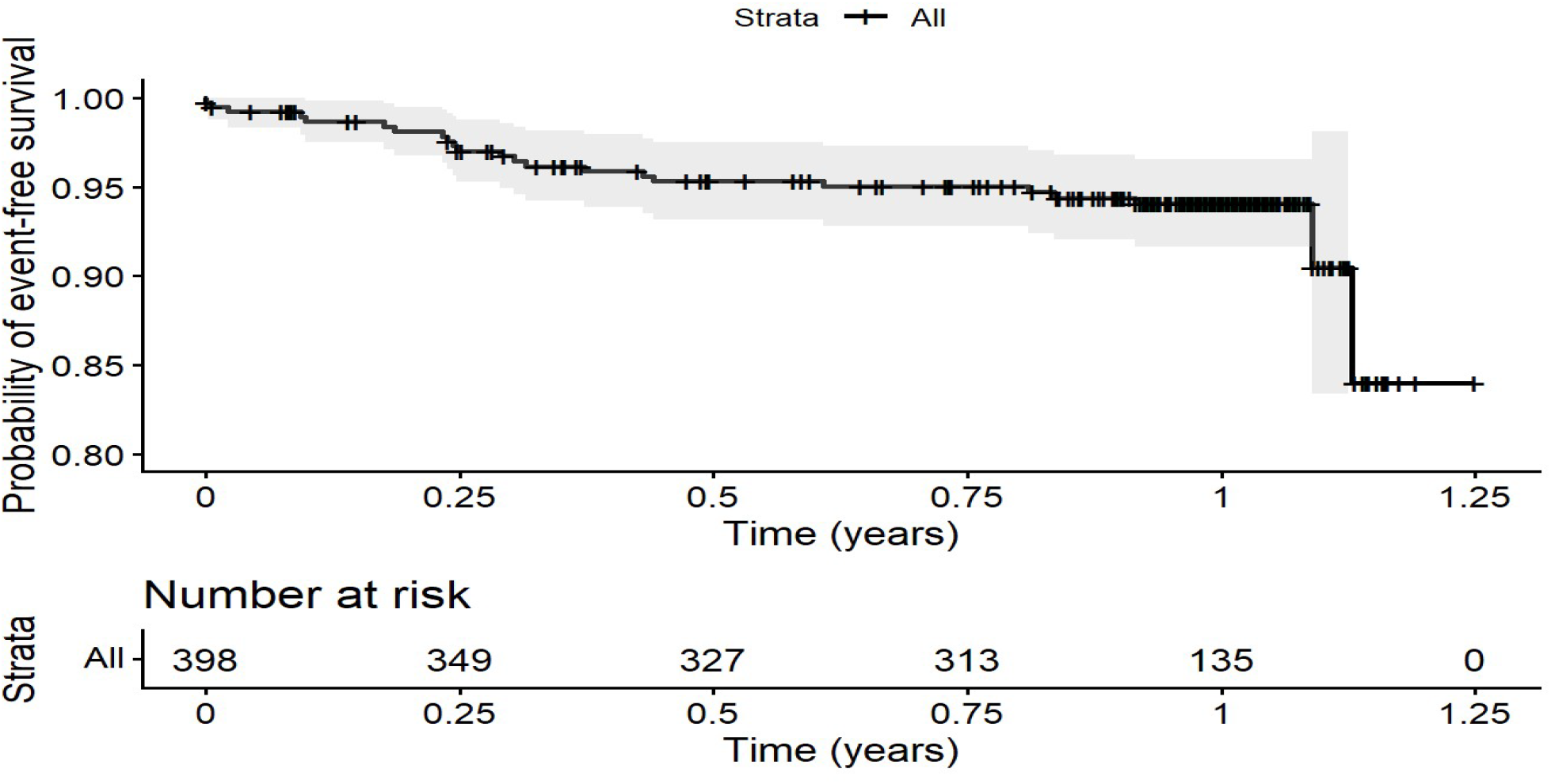
Event-free (Kaplan-Meier) for all-cause death, major bleeding events, and thromboembolic events (stroke/TIA).

The vast majority of patients (94%) didn’t experience any of the following treatment-related outcomes: all-cause death, major bleeding, or thromboembolic events **(freedom from events: Fig 2B)**. Outcome analysis demonstrated that major bleeding events and all-cause death increased in patients with higher risk scores of CHA_2_DS_2_-VASc **(Figure 3)**.

**Figure 3.**
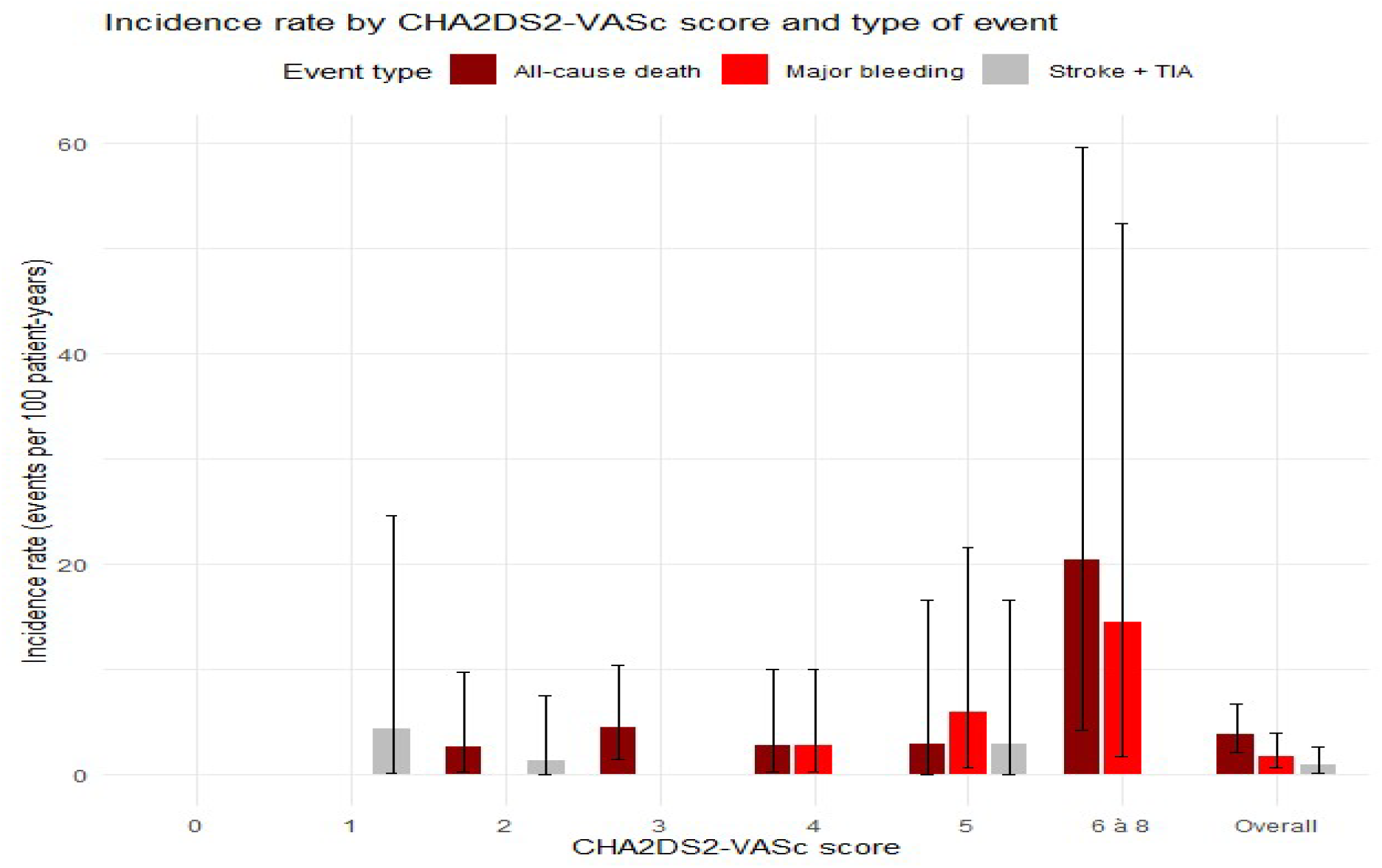
Outcomes as a function of CHA_2_DS_2_-VASc scores.

Treatment persistence was represented by 75,88% at the end of the study where 96 patients (24.12%) had at least one interruption of rivaroxaban treatment; most of the interruptions were due to patients lost to follow-up (36.4%), patient decisions (17.7%), death (12.7%), SAEs (5.5%), and AEs (2.7%). Other reasons (25.5%) included mainly the cost of the medication, which alone accounted for nearly half of the cases (46.4%).

## Discussion

Considering an estimated AF prevalence in the Algerian population of approximately 0.5%, and in the absence of studies conducted in Algeria regarding AF treatments, we carried out TROMBIX-DZ, the first real-world observational evaluation of rivaroxaban in Algerian patients with AF. Several studies were conducted previously in Europe, Canada, middle East, Africa, Latin America and Asia to evaluate the use of rivaroxaban in patients with AF in real-world settings. This study aimed to address the lack of regional data by enriching the literature with clinical details related to Algerian patients.

In this prospective observational study, the mean age was 70.5 ± 11.94 years which is close to other studies. Comparable proportions of patients with a CrCl <50 ml/min, common comorbidities with predominance of hypertension and diabetes were also observed. Additionally, CHA_2_DS_2_-VASc and HAS-BLED scores were mostly similar to previous studies **(6, 7, 8)**. Interestingly, a lower incidence of prior stroke/TIA was recorded in TROMBIX-DZ (8.8%), which could be explained by the high number of newly diagnosed patients.

The incidence proportion of treatment emergent SAEs and AEs were low in TROMBIX-DZ compared to other studies **(6, 7, 8)**. This might be due to the difference in medical system and the nature of the study. The rates of major bleeding were similar between TROMBIX-DZ and XANTUS (2.1 events per 100 patient-years) and comparable to XANTUS EL and XANAP (0.9 and 1.5 events per 100 patient-years). The same applied to Intracranial hemorrhages (0.3 vs. 0.4, 0.2 and 0.7, events/100 patient-years, respectively) and fatal bleeding (0.3 vs 0.2, 0.2 and 0.1 events/100 patient-years each). The rate of gastrointestinal bleeding was slightly higher (1.2 vs 0.9, 0.5 and 0.5 events/100 patient-years in XANTUS, XANAP, and XANTUS EL, respectively). A higher incidence rate of GI bleeding was also observed in NOAC study in patients using 20mg rivaroxaban (2.1%). It’s noteworthy that patient’s profile for the cases of major bleeding reveal two patients with CrCl below 50ml/min^2^ who were prescribed 20mg rivaroxaban where one of them experienced gastrointestinal bleeding. AEs increased during the course of the study; however, the vast majority of patients didn’t experience any treatment related adverse events (94%) which is similar to the values reported in the literature **(8)**. Rates of stroke were also similar between TROMBIX-DZ, XANTUS EL, and XANTUS (0.6, 0.6, and 0.7 events/100 patient-years, respectively). Overall stroke risk appeared comparable across patients despite differences in baseline characteristics.

The rate of all-cause death in our study was higher than previous studies, but is consistent with the global findings where all-cause mortality among people with AF is high particularly in low- and middle-income countries and largely linked to health-system and risk-factor patterns **(11)**. Incidence rate of treatment-emergent non-major bleeding was lower than previous studies. These findings emphasize on the safety of Trombix® in real-life clinical setting.

Drug persistence represents a major challenge in patients with AF, as discontinuation of anticoagulant therapy leaves patients inadequately protected against the risk of stroke. Persistence at one year in TROMBIX-DZ was slightly lower than previous studies **(6, 8)**. This could be due to the cost of the medication and its lack of reimbursement which influenced the continuity of follow-up in some patients.

Moreover, although our sample size is smaller than international cohort studies, it was calculated based on a defined precision level to ensure statistically accurate and clinically significant estimations for the observed incidence rates.

As the first study conducted with rivaroxaban in Algeria, it is also the only clinical study performed in the Maghreb region. This research aims to provide valuable prospective, real-world data on Algerian patients and reinforce the clinical data available for North Africa.

### Study limitations

Several limitations should be acknowledged. First, TROMBIX-DZ was an observational single-arm design, which may introduce bias related to patient evaluation, treatment and follow up. In addition to a potential of under-reporting especially for adverse event. Furthermore, in this study, there was no central adjudication committee responsible for adjudicating major bleeding, thromboembolic events, and all-cause mortality; the investigators reported the primary and secondary study outcomes which may result in reporting bias.

## Conclusion

TROMBIX-DZ represents the first cohort study in Algeria and the Maghreb region, describing the use of rivaroxaban for stroke prevention in an Algerian AF population. Our main findings are more aligned with those reported in XANTUS EL, this could be potentially due to the study focus on a comparable ethnic population (Middle eastern). Also, this pattern reinforces the external validity of our data and emphasize on the importance of real-world evidences. The results of this study are broadly consistent with those reported in various international real-world registries. Rates of treatment emergent adverse events including major bleeding were lower in TROMBIX-DZ which lines with a lower baseline bleeding risk and highlights the safety and efficacy of rivaroxaban on Algerian patients with AF.

Notwithstanding these similarities, the observed discrepancies might be attributed to different demographics, socioeconomic factors as well as a distinct healthcare system.

The collected data along with the obtained results provide valuable information on patient profile, therapeutic approach, safety, and the efficacy of rivaroxaban. thus, contribute to enriching knowledge about its use in real life settings and deliver an evidence-based medicine support that position its role in local clinical guidelines.

In conclusion, this observational study provides comprehensive and complementary data on the use of rivaroxaban in real-life clinical practice in the Maghreb region.

## Data Availability

Data statement
The datasets used or analyzed during the current study are available from the corresponding author upon reasonable request.

## Acknowledgements

We would like to thank all the patients who participated in the study, as well as the investigators and their associated teams Dr. Yacine Tir, Dr. Sawsen Sahnoune, Dr Amira Tir, Dr. Sarah Chebah, Dr Soraya Bachtarzi, Dr. Mohamed Mehdi Baouni, and Dr. Nawel Dahimene. We thank Professor Samira Abrouk for her rich contribution to the statistical analysis of this study. We acknowledge Professor Salam Bennouar for her valuable review of the manuscript

## Conflict of interest

ASMB, SL, SH, MA, IB and LN are employees of BEKER Laboratories. All others received funding as part of this study.

The CRO Clinica Group received funding for its contribution from BEKER Laboratories.

## Funding

This study was funded and sponsored by BEKER Laboratories. BEKER’s team participated in the design of the study, writing, and the decision to publish this article.

## Author’s contribution statement

ASMB, MA, developed this study’s protocol with input from the CRO “Clinica Group.” YA and NA validated the protocol. All clinical investigators (MSAM, NL, SB, AK, MFB, SB, TB, RO, ZK, BA, RH, MM, HMAL, MA, HE, NL, NN, YA, NN, GS and TF) were responsible for patients’ follow-up and acquisition of clinical data. Statistical analysis was conducted by an external paid consultant. ASMB, SL, SH, and MA conducted results interpretation. SL wrote the first draft. SL and ASMB co-wrote and revised the final manuscript. ASMB, SL, SH, MA, IB, LN, YA and NA revised the manuscript. All the authors confirmed the final published version.

## Data statement

The datasets used or analyzed during the current study are available from the corresponding author upon reasonable request.

## Notes

### Clinical Protocols

https://clinicaltrials.gov/study/NCT06184204

### Author Declarations

The study was conducted following approval of the Ethical Committee (EC) of Issad Hassani University Hospital of Beni Messous (N 8 / CE/2023) and the Algerian Ministry of Industry and Pharmaceutical Production (MIPP) (012/BEK/OBS/MED/2023) in November 2023, according to the Declaration of Helsinki, Good Clinical Practice (ICH GCP E6) and current Algerian regulations.

## References

1. Van Gelder, I.C., Rienstra, M., Bunting, K.V., Casado-Arroyo, R., Caso, V., Crijns, H.J.G.M., De Potter, T.J.R., Dwight, J., Guasti, L., Hanke, T., Jaarsma, T., Lettino, M., Løchen, M.-L., Lumbers, R.T., Maesen, B., Mølgaard, I., Rosano, G.M.C., Sanders, P., Schnabel, R.B., Suwalski, P., Svennberg, E., Tamargo, J., Tica, O., Traykov, V., Tzeis, S., Kotecha, D., Group, E.S.D., Dagres, N., Rocca, B., Ahsan, S., Ameri, P., Arbelo, E., Bauer, A., Borger, M.A., Buccheri, S., Casadei, B., Chioncel, O., Dobrev, D., Fauchier, L., Gigante, B., Glikson, M., Hijazi, Z., Hindricks, G., Husser, D., Ibanez, B., James, S., Kaab, S., Kirchhof, P., Køber, L., Koskinas, K.C., Kumler, T., Lip, G.Y.H., Mandrola, J., Marx, N., Mcevoy, J.W., Mihaylova, B., Mindham, R., Muraru, D., Neubeck, L., Nielsen, J.C., Oldgren, J., Paciaroni, M., Pasquet, A.A., Prescott, E., Rega, F., Rossello, F.J., Rucinski, M., Salzberg, S.P., Schulman, S., Sommer, P., Svendsen, J.H., ten Berg, J.M., Ten Cate, H., Vaartjes, I., Vrints, C.J., Witkowski, A., Zeppenfeld, K., Simoni, L., Kichou, B., Sisakian, H.S., Scherr, D., Cools, F., Smajić, E., Shalganov, T., Manola, S., Avraamides, P., Taborsky, M., Brandes, A., El-Damaty, A.M., Kampus, P., Raatikainen, P., Garcia, R., Etsadashvili, K., Eckardt, L., Kallergis, E., Gellér, L., Guðmundsson, K., Lyne, J., Marai, I., Colivicchi, F., Abdrakhmanov, A.S., Bytyci, I., Kerimkulova, A., Kupics, K., Refaat, M., Bheleel, O.A., Barysienė, J., Leitz, P., Sammut, M.A., Grosu, A., Pavlovic, N., Moustaghfir, A., Yap, S.-C., Taleski, J., Fink, T., Kazmierczak, J., Sanfins, V.M., Cozma, D., Zavatta, M., Kovačević, D.V., Hlivak, P., Zupan, I., Calvo, D., Björkenheim, A., Kühne, M., Ouali, S., Demircan, S., Sychov, O.S., Ng, A., Kuchkarov, H., n.d. 2024 ESC Guidelines for the management of atrial fibrillation developed in collaboration with the European Association for Cardio-Thoracic Surgery (EACTS): Developed by the task force for the management of atrial fibrillation of the European Society of Cardiology (ESC), with the special contribution of the European Heart Rhythm Association (EHRA) of the ESC. Endorsed by the European Stroke Organisation (ESO).

2. Tan, S.C.W., Tang, M.-L., Chu, H., Zhao, Y.-T., Weng, C., 2025. Trends in Global Burden and Socioeconomic Profiles of Atrial Fibrillation and Atrial Flutter: Insights from the Global Burden of Disease Study 2021. CJC Open 7, 247–258. 10.1016/j.cjco.2024.11.017

3. HS_PREV_AFD_STD_100K_T_R,Prevalence of atrial fibrillation, Both [WWW Document], n.d. URL https://eatlas.escardio.org/Data/Cardiovascular-disease-morbidity/hs_prev_afd_std_100k_t_r-prevalence-of-atrial-fibrillation-both (accessed 2.12.26).

4. Patel, M.R., Mahaffey, K.W., Garg, J., Pan, G., Singer, D.E., Hacke, W., Breithardt, G., Halperin, J.L., Hankey, G.J., Piccini, J.P., Becker, R.C., Nessel, C.C., Paolini, J.F., Berkowitz, S.D., Fox, K.A.A., Califf, R.M., null, null, 2011. Rivaroxaban versus Warfarin in Nonvalvular Atrial Fibrillation. New England Journal of Medicine 365, 883–891. 10.1056/NEJMoa1009638

5. Pengo, V., Denas, G., 2018. Optimizing quality care for the oral vitamin K antagonists (VKAs). Hematology Am Soc Hematol Educ Program 2018, 332–338. 10.1182/asheducation-2018.1.332

6. Martínez, C.A.A., Lanas, F., Radaideh, G., Kharabsheh, S.M., Lambelet, M., Viaud, M.A.L., Ziadeh, N.S., Turpie, A.G.G., 2018. XANTUS-EL: A real-world, prospective, observational study of patients treated with rivaroxaban for stroke prevention in atrial fibrillation in Eastern Europe, Middle East, Africa and Latin America. The Egyptian Heart Journal 70, 307–313. 10.1016/j.ehj.2018.09.002

7. Kim, Y.-H., Shim, J., Tsai, C.-T., Wang, C.-C., Vilela, G., Muengtaweepongsa, S., Kurniawan, M., Maskon, O., Li Fern, H., Nguyen, T.H., Thanachartwet, T., Sim, K., Camm, A.J., Investigators, the X., 2018. XANAP: A real-world, prospective, observational study of patients treated with rivaroxaban for stroke prevention in atrial fibrillation in Asia. Journal of Arrhythmia 34, 418–427. 10.1002/joa3.12073

8. Camm, A.J., Investigators, the X., Amarenco, P., Investigators, the X., Haas, S., Investigators, the X., Hess, S., Investigators, the X., Kirchhof, P., Investigators, the X., Kuhls, S., Investigators, the X., van Eickels, M., Investigators, the X., Turpie, A.G.G., Investigators, the X., n.d. XANTUS: a real-world, prospective, observational study of patients treated with rivaroxaban for stroke prevention in atrial fibrillation.

9. Kocabaş, U., Ergin, I., Sönmez, S.C., Yavuz, V., Murat, S., Özdemir, I.H., Genç, Ö., Tüner, H., Meriç, B.K., Aslan, O., Dal, A., Taşkın, U., Şen, T., İbişoğlu, E., Erdoğan, A., Özgeyik, M., Demir, M., Urgun, Ö.D., Doğduş, M., Çakal, S., Çayırlı, S., Güler, A., Karabulut, D., Dalgıç, O., Murat, B., Karabulut, U., Öztekin, G.M.Y., Biter, H.İ., Sinan, Ü.Y., Barış, V.Ö., Kaplan, M., Altın, C., Kıvrak, T., 2025. Incidence and Predictors of Clinical Outcomes in Real-Life Patients With Atrial Fibrillation Treated With Oral Factor Xa Inhibitors: The Follow-Up Results of the ANATOLIA-AF Study. Clin Cardiol 48, e70088. 10.1002/clc.70088

10. Gedikli, Ö., Altay, S., Ünlü, S., Çakmak, H.A., Aşkın, L., Yanık, A., Beşli, F., Sinan, Ü.Y., Canpolat, U., Şahin, M., Pehlivanoğlu, S., 2021. Real-life data of major and minor bleeding events with direct oral anticoagulants in the one-year follow-up period: The NOAC-TURK study. Anatol J Cardiol 25, 196–204. 10.5152/AnatolJCardiol.2021.57635

11. Santos IS, Goulart AC, Olmos RD, et al. Atrial fibrillation in low- and middle-income countries: a narrative review. European Heart Journal Supplements : Journal of the European Society of Cardiology. 2020;22(Suppl O):O61. doi:10.1093/eurheartj/suaa181

